# Comparative Evaluation of 19 Reverse Transcription Loop-Mediated Isothermal Amplification Assays for Detection of SARS-CoV-2

**DOI:** 10.1101/2020.07.22.20159525

**Authors:** Yajuan Dong, Xiuming Wu, Shenwei Li, Renfei Lu, Zhenzhou Wan, Jianru Qin, Guoying Yu, Xia Jin, Chiyu Zhang

## Abstract

Coronavirus disease 2019 (COVID-19) caused by SARS-CoV-2 has caused a global pandemics. To facilitate the detection of SARS-CoV-2 infection, various RT-LAMP assays using 19 sets of primers had been developed, but never been compared. We performed comparative evaluation of the 19 sets of primers using 4 RNA standards and 29 clinical samples from COVID-19 patients. Six of 15 sets of primers were firstly identified to have faster amplification when tested with four RNA standards, and were further subjected to parallel comparison with the remaining four primer sets using 29 clinical samples. Among these 10 primer sets, Set-4 had the highest positive detection rate of SARS-CoV-2 (82.8%), followed by Set-10, Set-11, Set-13 and Set-14 (75.9%), and Set-14 showed the fastest amplification speed (< 8.5 minutes), followed by Set-17 (< 12.5 minutes). Based on the overall detection performance, Set-4, Set-10, Set-11, Set-13, Set-14 and Set-17 that target *Nsp3, S, S, E, N* and *N* gene regions of SARS-CoV-2, respectively, are determined to be better than the other primer sets. Two RT-LAMP assays with the Set-14 primers in combination with any one of four other primer sets (Set-4, Set-10, Set-11, and Set-13) are recommended to be used in the COVID-19 surveillance.

## Introduction

Coronavirus disease 2019 (COVID-19), caused by the newly discovered coronavirus SARS-CoV-2 ^1,2^, is rapidly spreading throughout the world, posing a huge challenge to global public health security. As of 1 June, 2020, it has infected over 6 million people, and resulted in at least 376,320 deaths globally. In the absence of effective antiviral drugs or efficacious vaccines, early diagnosis of SARS-CoV-2 infection is essential for the containment of COVID-19 ^3,4^, without which it is impossible to timely implement intervention and quarantine measures, and difficult to track contacts in order to limit virus spread.

Nucleic acid testing of various approaches are widely used as the primary tool for diagnosing COVID-19 ^3,4^. Among them, real-time quantitative PCR (RT-qPCR) methods have been set as the gold standard for laboratory confirmation of SARS-CoV-2 infection because of their proven track record as being the most robust technology in molecular diagnostics ^4-6^. However, the RT-qPCR assay relies on sophisticated facilities with reliable supply of electricity and well-trained personnel in large general hospitals and health care facilities, or government labs (such as CDC), and it is relatively time-consuming (about 1.5-2 hrs). These limit its capacity in point-of-care settings. Moreover, visiting a clinical setting for testing increases the risk of spreading the virus. Therefore, an alternative, fast, simple, and sensitive point-of-care testing (POCT) is highly needed to facilitate the detection of SARS-CoV-2 infection in resource-limited settings ^3,7^.

Loop-mediated isothermal amplification (LAMP) is a promising POCT method with high sensitivity, specificity, and rapidity, and it is easy-to-use ^8^. To overcome the limitation of RT-qPCR assay, a number of RT-LAMP assays using at least 19 sets of different primers had been developed in the last few months for the detection of SARS-CoV-2 ^9-19^. Although these assays had proven sensitive and effective for the detection of SARS-CoV-2, how do they compare with each other have not been evaluated. In this study, we compared all 19 sets of SARS-CoV-2-specific RT-LAMP primers using the mismatch-tolerant LAMP system that is faster and more sensitive than the conventional ones ^20,21^, and screened the high-efficiency RT-LAMP assays for use in the detection of field samples.

## Results

### Strategy for the comparative evaluation

There were 19 sets of SARS-CoV-2 RT-LAMP primers available for the evaluation. Among these primers, 2 sets were designed for binding to the *Nsp3* (non-structural proteins), 5 for *RdRp* (RNA-dependent RNA polymerase), 2 for *E* (envelope protein) and 2 for *N* (nucleocapsid protein) gene regions of SARS-CoV-2 (Fig. 1). These regions are highly conserved among SARS-CoV-2 and SARS-CoV, but distinct from five other human coronaviruses (MERS-CoV, OC43, 229E, NL63 and HKU1). Other 4 sets of primers were dispersed throughout the genome of SARS-CoV-2. The primers binding to the same target gene are adjacent to each other and cover genomic segments from 251 to 1954 bps. To minimize the consumption of clinical samples, and economize experimental efforts, we adopted a strategy that initiated by a preliminary evaluation of the primers binding to the four major genomic regions using *in vitro*-transcribed RNA standard, and followed by a further evaluation of preliminarily selected primers together with four sets of other primers using clinical RNA samples (Fig. 1).

**Figure 1.**
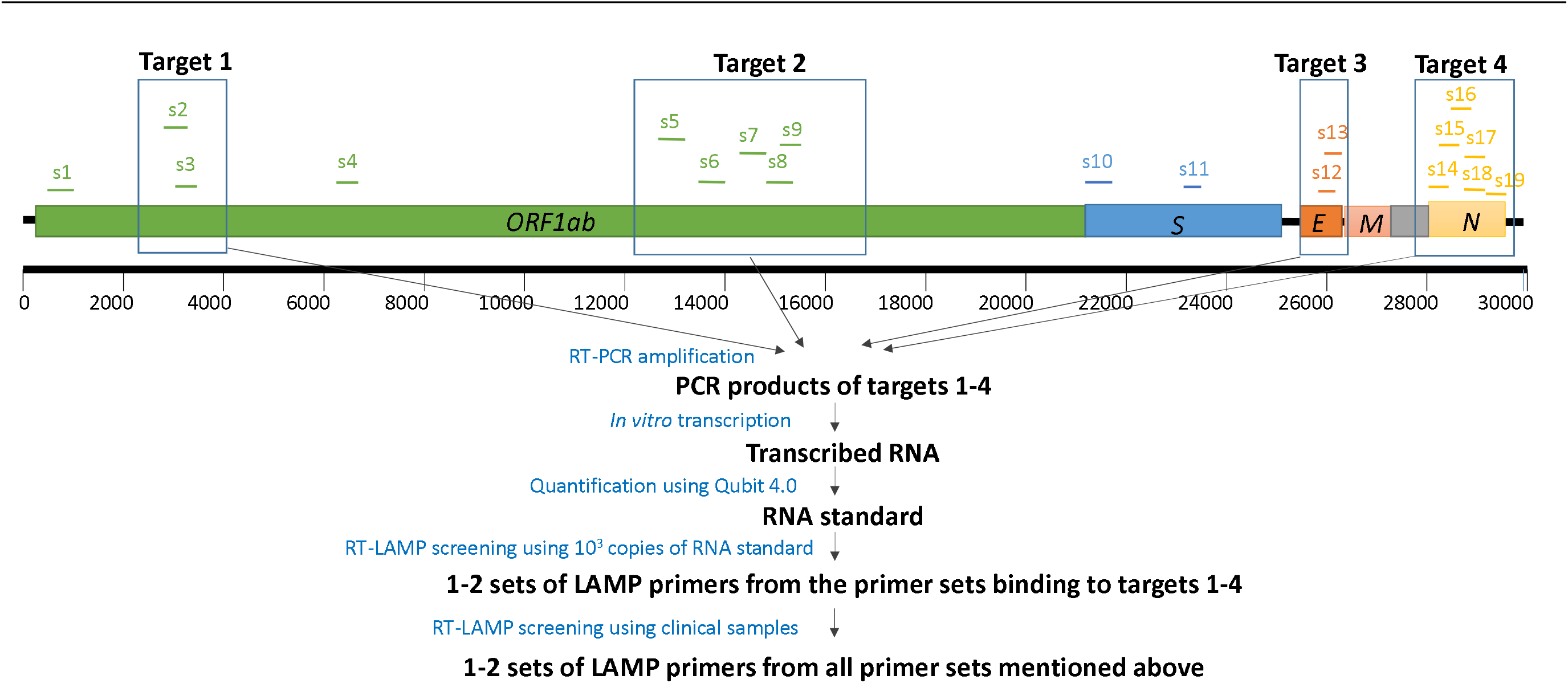
Genome location and evaluation strategy of 19 sets of SARS-CoV-2 RT-LAMP primers. The location of each primer set was detailed in Table 1. SD: standard deviation.

**Table 1.**
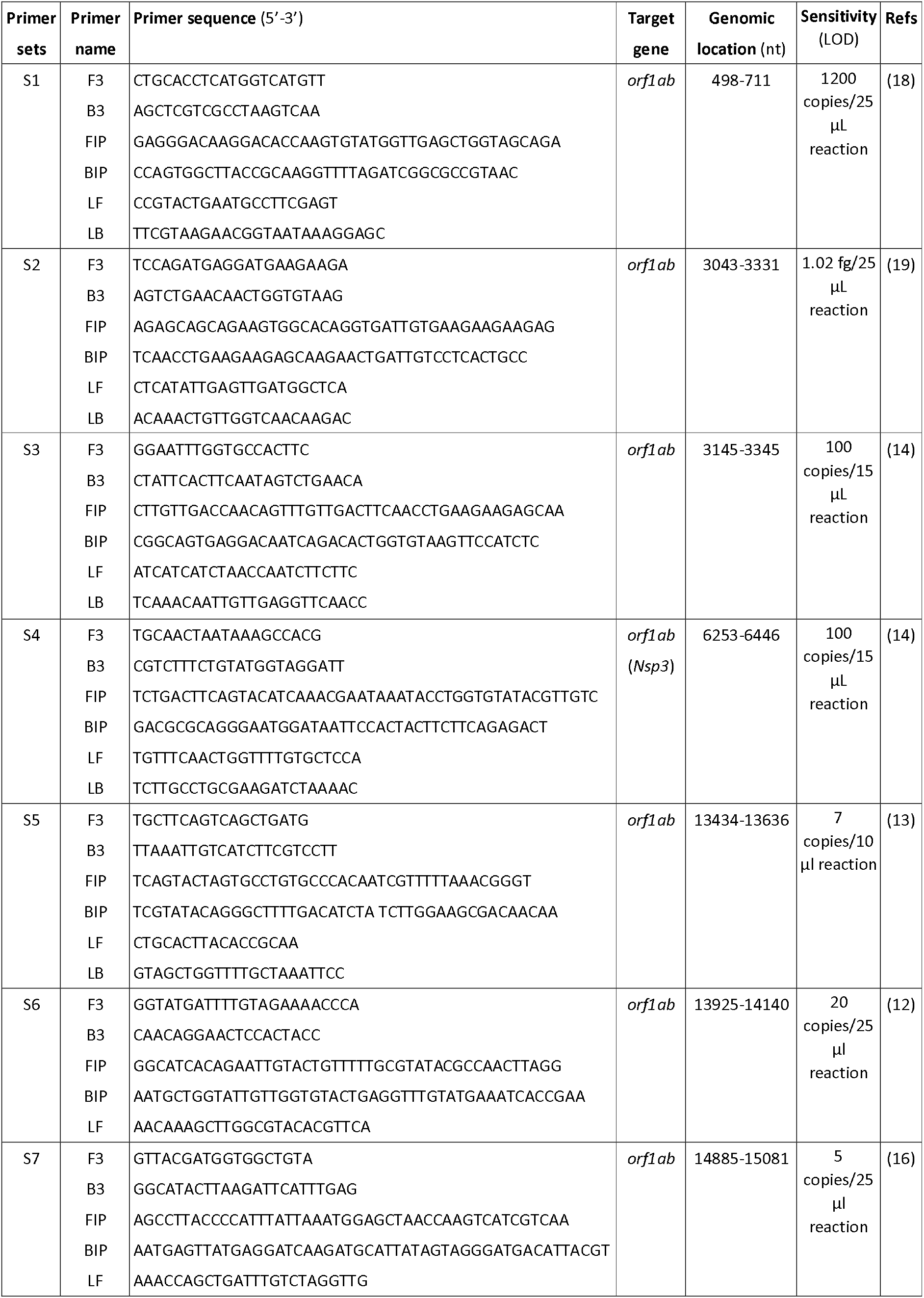
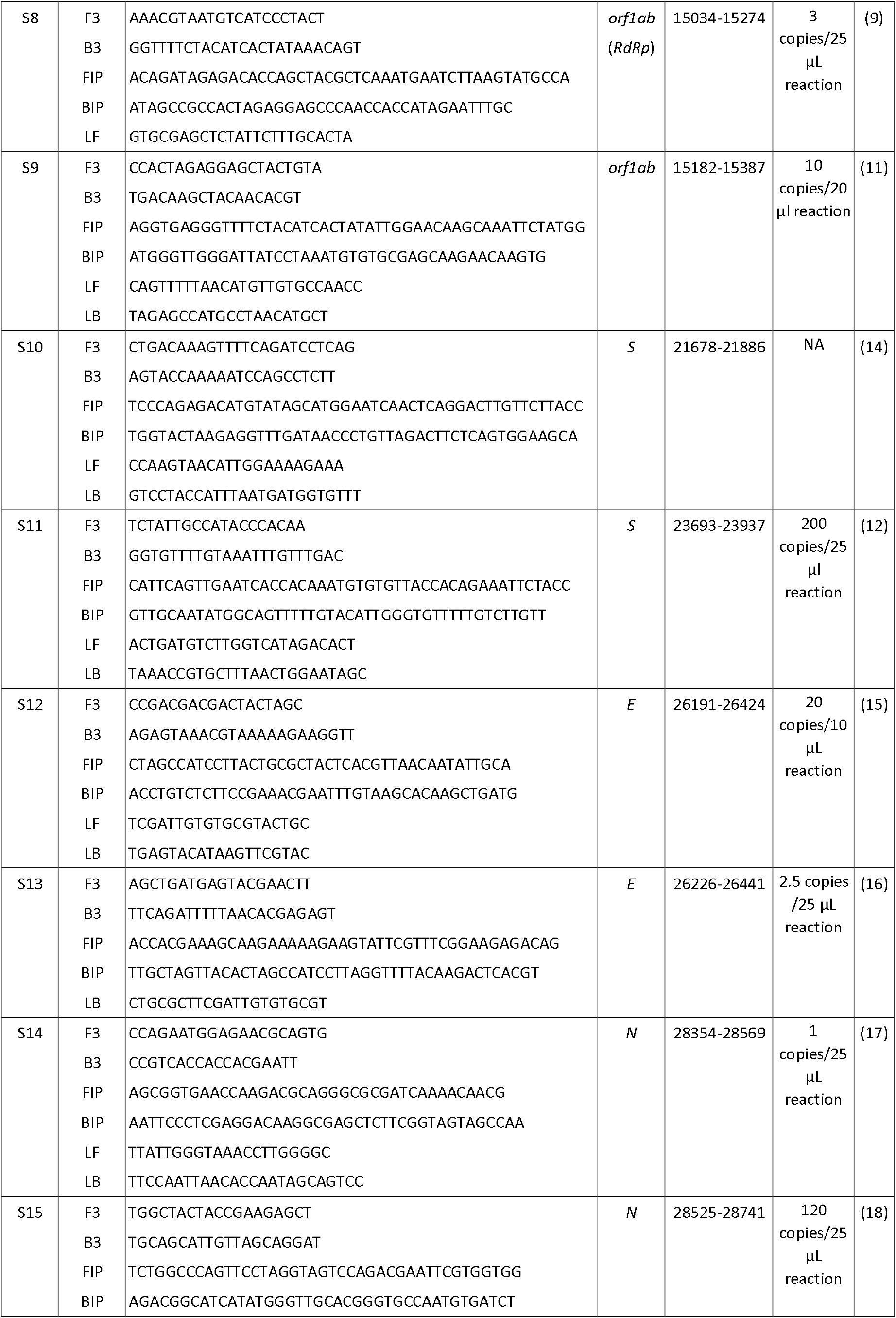
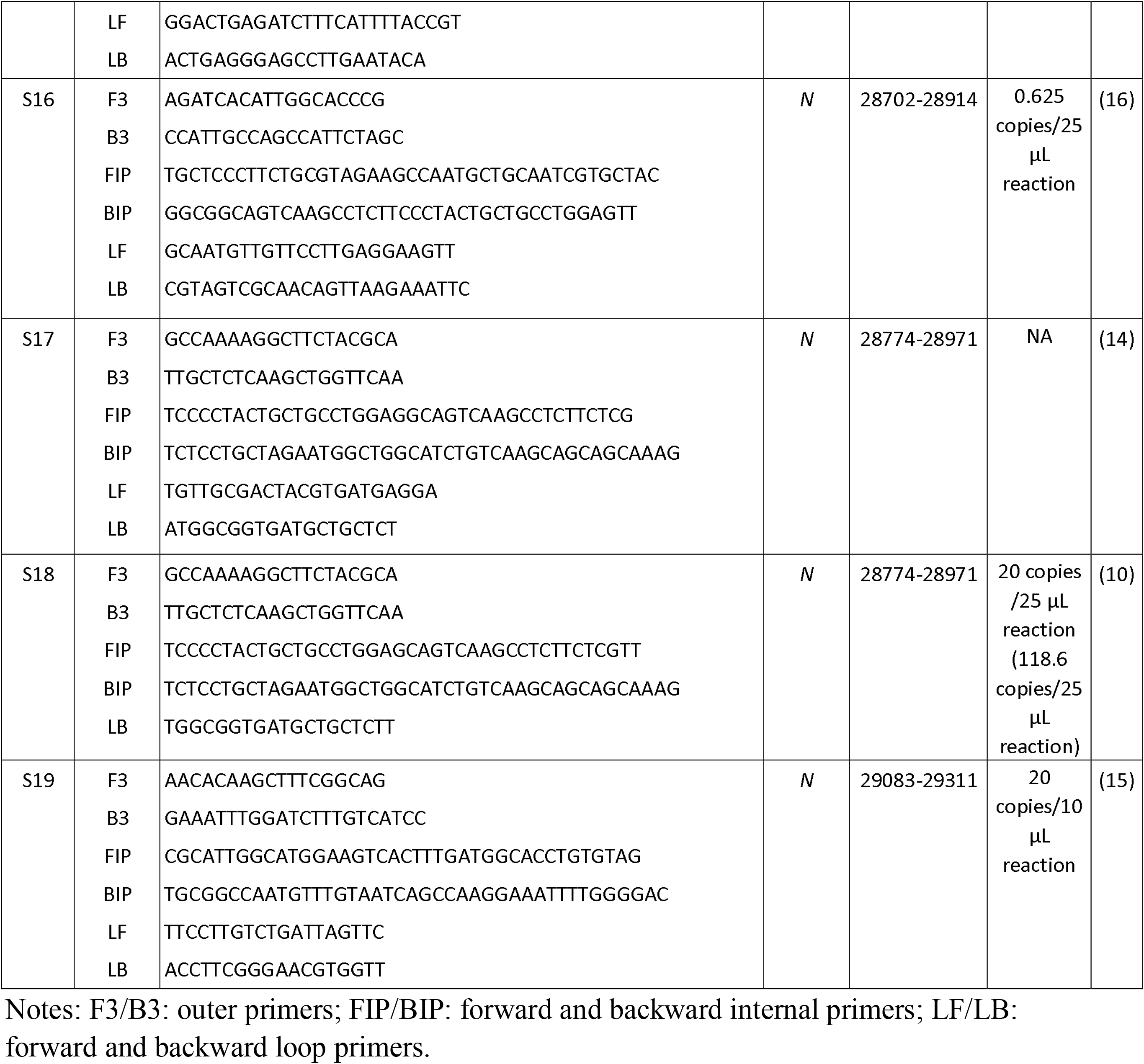
Information of 19 sets of RT-LAMP primers for the detection of SARS-CoV-2.

### Preliminary evaluation of primer sets

Using 3000 copies of *in vitro*-transcribed RNA standards of four gene segments of SARS-CoV-2, we assessed the amplification performance of 15 sets of RT-LAMP primers. Except for Set-3 that failed in amplification, all other primer sets generated amplification curves with Time threshold (Tt) of 7.5-15.9 minutes and reached the plateau phase within 20 minutes (Fig. 2). In particular, six sets of the primers showed faster amplification with 10 minutes less Tt values than other primer sets (Fig. 2), implying higher amplification sensitivity. The six sets (Set-2, Set-5, Set-13, Set-14, Set-17 and Set-18) of primers contain three that bind to *N* gene and another three that bind to *Nsp, RdRp*, and *E* genes, respectively. The six sets of primers were selected for further evaluation using clinical samples together with other four primer sets that bind to other genomic regions of the virus.

**Figure 2.**
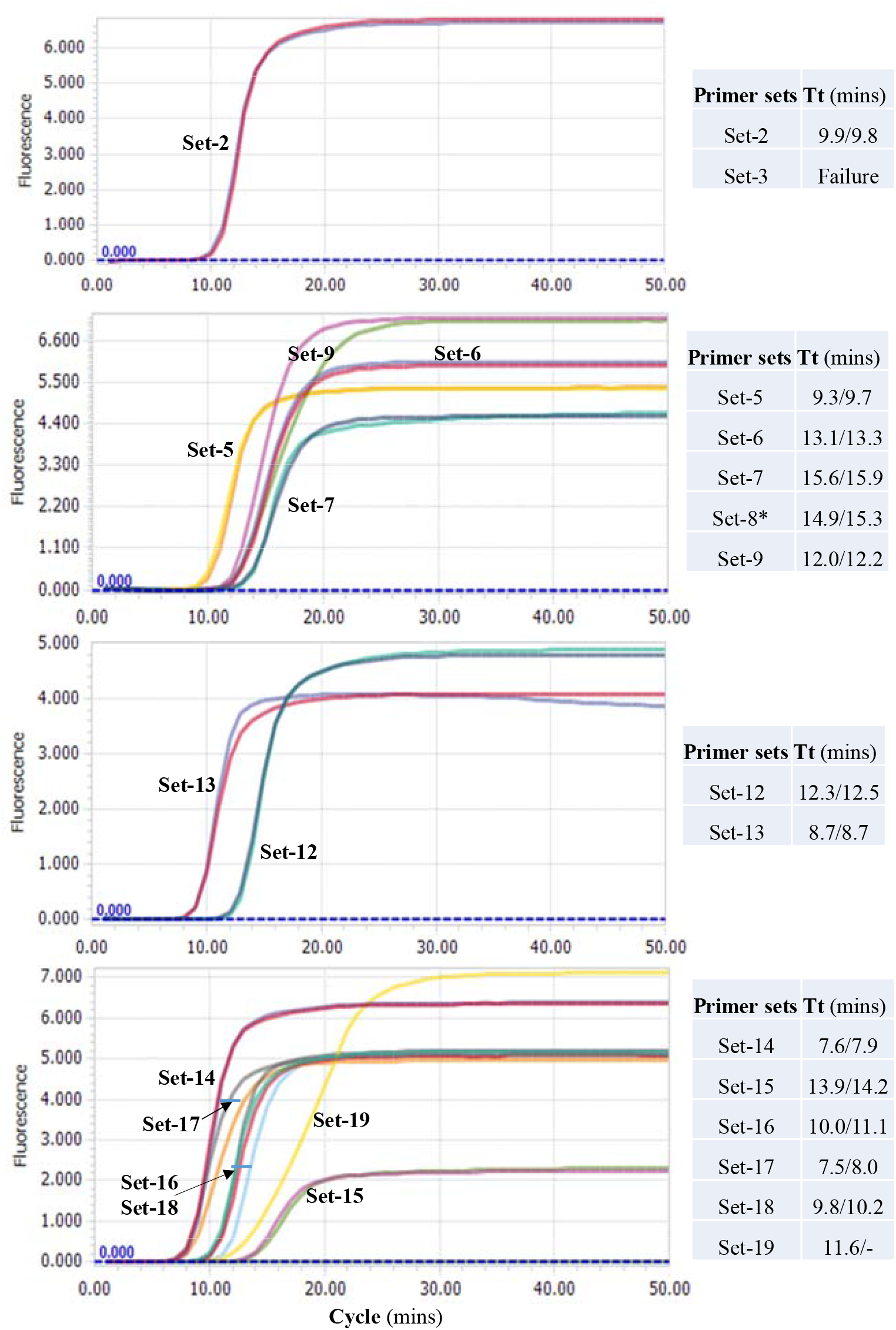
Comparison of performance of 15 RT-LAMP assays using RNA standards. The curves of non-template control (NTC) are not shown. * The Tt values of the Set-8 were obtained by another repeated comparative experiments with Set-5and Set-9, both of which showed a consistent trend, but slightly lower Tt values than those shown here.

### Comparative evaluation of ten primer sets using clinical samples

A total of 29 RNA samples extracted from COVID-19 patients were used at 4-fold dilutions. Except for one, all 29 RNA samples were detected as being SARS-CoV-2 positive by at least one of the primer sets. Nine samples were detected as positive by all ten sets of primers and almost all reactions (except for one with 49.5 minutes) had Tt values of less than 15.1 minutes, indicating a high viral load. The primer Set-4 detected 24 positive samples, showing the highest positive detection rate (82.8%), followed by Set-10, Set-11, Set-13 and Set-17 that all detected 22 positive samples (75.9%) (Fig. 3A). Two primer sets, Set-1 and Set-18, had the lowest positive detection rates of 44.8% and 62.1%, respectively, and thus were excluded in the subsequent analyses. Comparison showed that the primer Set-14 had the lowest mean Tt values of less than 8.4 minutes, followed by Set-10, Set-11 and Set-13 that had mean Tt values of 11.1-11.5 minutes (Fig. 3A). These four fast-amplification sets of primers also have small standard deviations (SD) of 1.7-2.9, indicating that the RT-LAMP with these four primer sets are relatively more stable and faster than the other 15 sets. As expected, the primer Set-14 is the most efficient one that generated the fastest (the lowest Tt value) and the second fastest amplification in 14 and 7 samples, followed by Set-17 which is the fastest in 6 samples and second best in 9 samples, demonstrating these two primer sets had the best performance.

**Figure 3.**
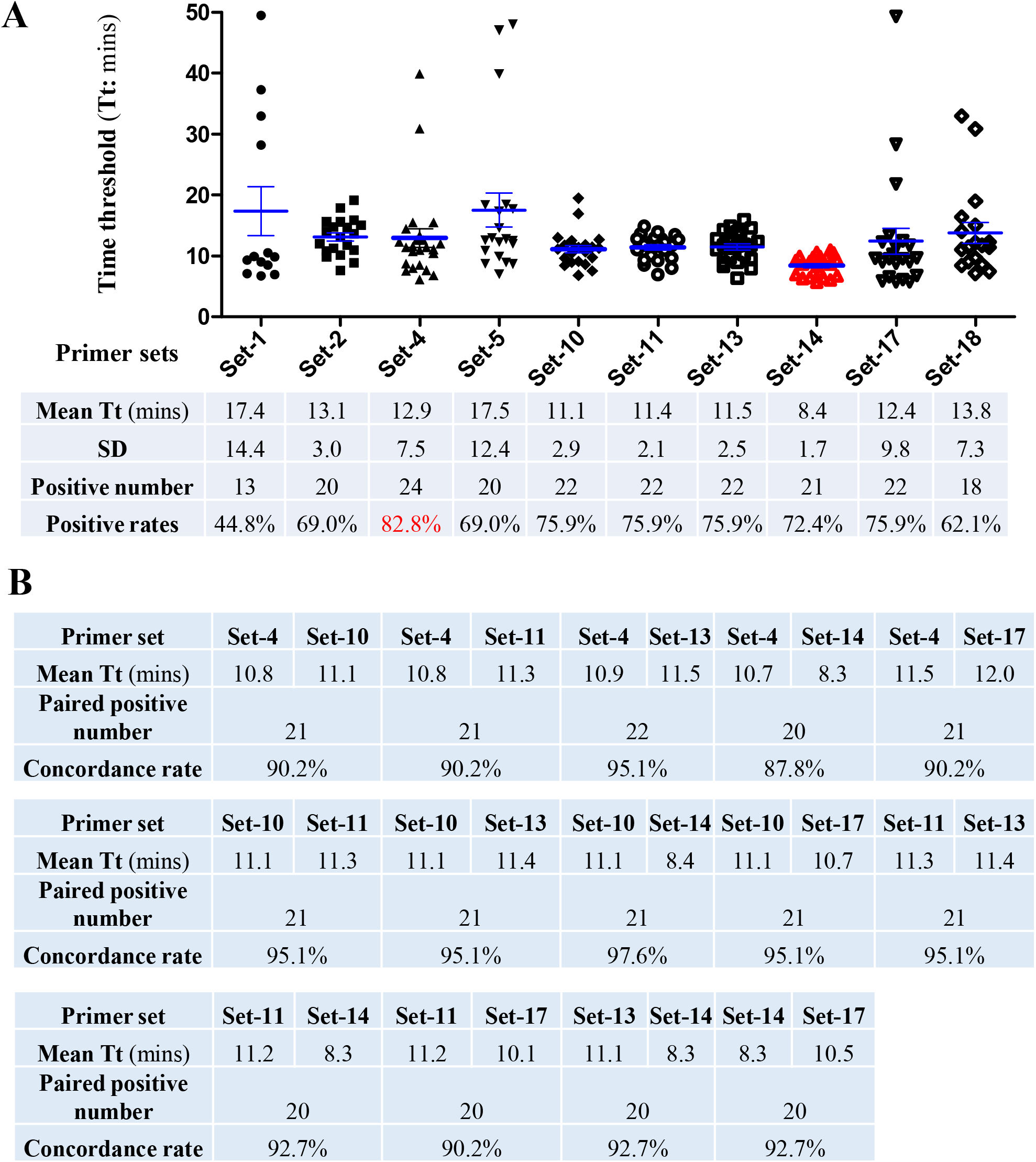
Comparison of performance of 10 selected RT-LAMP primer sets using 41 clinical RNA samples. A. Positive rates and Tt values of 10 selected RT-LAMP assays. B. Paired comparison of Tt values of the primers Set-4, Set-10, Set-11, Set-13, Set-14 and Set-17.

Because of their relatively high positive detection rate and lower Tt values, six primer sets including Set-4, Set-10, Set-11, Set-13, Set-14 and Set-17 were subjected to further pairwise comparison. The comparison showed that any two sets of these primers had high concordance performance (87.8-97.6%) for 41 clinical RNA samples (including 29 positive and 12 negative for SARS-CoV-2) (Fig. 3B). All the six primer sets had high amplification efficiency with mean Tt values of less than 12 minutes (Fig. 3B). In particular, Set-14 had faster amplification than the other five sets of primers (8.3-8.4 vs. 10.5-11.2 minutes).

### Specficity evaluation of four optimal primer sets based on sequence alignment

The specificity of these primer sets had been reported in previous studies ^9-19^. In this evaluation, all ten primer sets did not generate amplification for all 12 COVID-19 negative RNA samples. To further examine the specificity of six recommended primer sets (Set-4, Set-10, Set-11, Set-13, Set-14 and Set-17) to other human coronaviruses, we performed sequence alignment analyses. SARS-CoV-2 shared 79.5% genomic homology with SARS-CoV ^1,2^, indicating a relatively high sequence identity; but it was largely distinct from MERS-CoV and other four human coronaviruses (Supplementary Fig. S1). In particular, several primers of Set-4, Set-10 and Set-17 correspond to gaps or insertions of the genomes of MERS-CoV and other four common human coronaviruses OC43, 229E, NL63 and HKU1. These results implied that these six sets of primers were unable to bind to the genomes of MERS-CoV and four common human coronaviruses, therefore more specific for SARS-CoV-2. However, because of high sequence identity and the use of mismatch-tolerant RT-LAMP system that allows the presence of few mismatched bases between primers and templates, the SARS-CoV-2 RT-LAMP assays may generate a cross-amplification of SARS-CoV.

## Discussion

SARS-CoV-2 transmission mainly occurs in the early and progressive stages of COVID-19 disease during which the patients and virus carriers have higher viral load than that in recovery stage ^22-24^, and are generally more infectious. To contain the spread of the virus, early diagnosis is essential ^3,4^. It helps to trigger timely intervention (e.g. quarantine, lockdown, and contact tracing), and facilitates to optimize clinical management. It is clear that serological assays are not suitable for this purpose, because detectable antibodies always appear several days after infection. Therefore, viral RNA testing is the primary method for early diagnostics of COVID-19. Despite being the most robust diagnostic tests, RT-qPCR-based assays are more centralized in core facilities, and they are not amenable for large-scale monitoring for asymptomatic and pre-symptomatic virus carriers in point-of-care settings (e.g. community and home). Therefore, community- and/or home-based nucleic acid assays that allow individuals to test in the community, at home, or other point-of-care sites without having to visit hospitals are convenient tools for the detection of SARS-CoV-2 infection by the general public ^3,7^.

RT-LAMP assays are such needed tools ^8,20,21^. In fact, various LAMP assays have been developed that included at least 19 sets of primers targeting different genomic regions of SARS-CoV-2, with reported high sensitivity of detection ranging from 0.625 to 1200 copies per 25 µL reaction ^9-19^. However, these primers are never formally evaluated with clinical samples. The sensitivity and performance of a RT-LAMP assay are mainly determined by the primers set, because other components of the reaction system are optimized and stable. Therefore, assessing the optimal RT-LAMP primer sets for the detection of SARS-CoV-2 infection are important for the selection of best assay format to use for large field screening of COVID-19 patients.

Recently, the reaction system of RT-LAMP was further optimized to have higher sensitivity and faster amplification speed, even allowing the presence of few mismatched bases between primer and templates in a mismatch-tolerant version ^20,21^. Using this new version, we assessed 19 sets of SARS-CoV-2 RT-LAMP primers. Six sets of primers with faster amplification speed were firstly selected from 15 sets of primers using 4 RNA standards, and then tested with other 4 primer sets using 41 clinical samples. Eight sets of primers showed either comparable or better performance than the other 2 sets of primers (Set-1 and Set-18) as determined by positive detection rate (>69.0%). Of the 8 sets of primers, six were further selected based on high positive detection rate and/or overall faster amplification speed (with mean Tt of less than 13 minutes). The six primer sets are Set-4, Set-10, Set-11, Set-13, Set-14 and Set-17 that correspond to *Nsp3, S, S, E, N*, and *N* genes of SARS-CoV-2, respectively.

Among these primer sets, the *N* gene-based RT-LAMP assays (Set-14 and Set-17) had the fastest amplification speed, followed by *S* and *E* gene-based assay (Set-10, Set-11 and Set-13). This result suggested that the *N* gene-based RT-LAMP assay is more sensitive in detecting SARS-CoV-2 than that based on other genes, consisting with results of RT-qPCR assays ^5^. Interestingly, previous studies showed that the sensitivities of Set-4 and Set-11 were more than 100 copies per 25 µL reaction ^12,14^, not much higher than our assay. In this study, both primer sets generated comparable performance with highly sensitive primers Set-13 and Set-14 (less than 3 copies per 25 µL reaction) ^16,17^. In addition, two of our previous reported primers, Set-8 and Set-18, exhibited high sensitivities of 3-20 copies per 25 µL reaction and good performance in the detection of clinical samples under the mismatch-tolerant reaction condition ^9,10^, but they did not show better performance than other nine primer sets in this study. A reason might be that the use of the mismatch-tolerant reaction system generally improved the amplification efficiency of the primers reported by other groups ^20^.

The analyzed primer sets showed high specificity in that they did not amplify any SARS-CoV-2 negative clinical samples. Sequence alignment analyses further supported that the six sets of optimal primers had good specificity to SARS-CoV-2, albeit they might generate non-specific amplification for SARS-CoV due to a high degree of sequence identity. However, given the lethal nature of both SARS-CoV-2 and SARS-CoV ^25^, a non-specific positive result for SARS-CoV might also be of clinical importance.

Two nucleic acid assays targeting different genes are suggested to be used in the detection of SARS-CoV-2 to avoid potential false-negative results ^5^. Based on comparable performances, any two of the six optimal primer sets (Set-4, Set-10, Set-11, Set-13, Set-14 and Set-17) are recommend to be used in the detection of SARS-CoV-2. However, simultaneous use of Set-10 and Set-11, or Set-14 and Set-17 should be avoided because the former two sets target *S* gene and the latter two sets target *N* gene. In addition, because of its very fast amplification speed, Set-14 is strongly encouraged to be preferentially selected for the diagnosis of COVID-19 patients. Apart from the six recommend primer sets, other primers such as Set-2 and Set-5 also had good performance, and can also be used in the monitoring of COVID-19 infections.

Another advantage of our version of the RT-LAMP assay is that the results are easily visualized with a pH-sensitive indicator dye (e.g. cresol red and neutral red) ^26^. Moreover, a combination of a nucleic acid extraction-free protocol and a master RT-LAMP mix containing all reagents (enzymes, primers, magnesium, nucleotides, dye and additives), except for the template, enables the development of a simple kit that can be used at home, or a community-based diagnosis center for the detection of COVID-19 infection ^3,27^.

In summary, we evaluated and selected six optimal primer sets from 19 sets of SARS-CoV-2 RT-LAMP primers through a comparative evaluation with clinical RNA samples from COVID-19 patients. Two RT-LAMP assays with the Set-14 primers and any one of the other four primer sets (Set-4, Set-10, Set-11 and Set-13) are strongly recommended to be used in the COVID-19 surveillance to facilitate the early finding of asymptomatic and pre-symptomatic virus carriers in clinical and point-of-care settings, and the monitoring of environmental samples in the field.

## Materials and Methods

### Ethics Statement

The study was approved by Nantong Third Hospital Ethics Committee (E2020002: 3 February 2020). All experiments were performed in accordance with relevant guidelines and regulations. Written informed consents were obtained from each of the involved patients.

### Preparation of RNA standard

To prepare RNA standard, four SARS-CoV-2 genomic segments (2720-3620 nt, 13403-15502 nt, 25901-26700 nt and 28274-29533 nt in Wuhan-Hu-1, GenBank: MN908947.3) were amplified from previously confirmed positive RNA sample with T7-promoter-containing primers (Supplementary Table S1). RNA standard was generated by *in vitro* transcription, and quantitated by Qubit® 4.0 Fluorometer (Thermo Fisher Scientific, USA). Copy number of RNA standard was estimated using the formula: RNA copies/ml = [RNA concentration (g/μL)/(nt transcript length × 340)] × 6.022 × 10^23^.

### RNA samples of COVID-19 patients

A total of 29 RNA samples were obtained from COVID-19 patients described in our previous studies ^9,10^. In brief, RNA was extracted from 300 μL throat swabs of COVID-19 patients using RNA extraction Kit (Liferiver, Shanghai) and eluted in 90 μL nuclease-free water. After screening and confirmation tests, the remaining RNA samples were stored at − 80 °C. When used for RT-LAMP assays, the stored SARS-CoV-2 positive RNA samples as confirmed by RT-qPCR assay were thawed and 4-fold diluted. In addition, 12 SARS-CoV-2 negative clinical RNA samples were used as controls..

### RT-LAMP Assay

To assess the performance of 19 sets of RT-LAMP primers in the detection of SARS-CoV-2, an optimized mismatch-tolerant RT-LAMP method that has higher sensitivity and faster amplification speed than the conventional ones was used. A 25 µL RT-LAMP reaction mixture was prepared with 1x isothermal amplification buffer, 6 mM MgSO4, 1.4 mM dNTPs, 8 units of WarmStart Bst 2.0 DNA polymerase, 7.5 units of WarmStartR RT, 0.15 unit of Q5 High-Fidelity DNA Polymerase, 0.2 μM each of primers of F3 and B3, 1.6 μM each of primers of FIP and BIP, 0.4 μM each of loop primer LF and/or LB, and 0.4 mM SYTO 9 (Life technologies, Carlsbad, CA, United States). The enzymes were all purchased from New England Biolabs (Beverly, MA, United States). In general, 3 μL of RNA standard or samples were added into each RT-LAMP reaction. The reaction was run at 63 □ for 50 minutes with real-time monitoring by the LightCycler 96 real-time PCR System (Roche Diagnostics, Mannheim, Germany).

## Data Availability

The all datas from the study have been uploaded.

## Acknowledgments

We thank the grants from the National Science and Technology Major Project of China (2019YFC1200603, 2017ZX10103009-002 and 2018ZX10711001) for financial support.

## Author Contributions

C.Z. conceived and designed the study, wrote the manuscript, and supervised the project. Y.D. and X.W. performed the experiments. S.L, R.L and Z.W collected and screened clinical samples. C.Z., X.W., Y.D., and J.Q. analyzed the data. C.Z., G. Y. and X. J. interpreted the results. X.J. contributed to critical revision of the manuscript.

## Authors’ Disclosures or Potential Conflicts of Interest

The authors report no conflicts of interest in this work.

